# Effectiveness of Mobilisation with Movement (MWM) along with Usual Care for Knee Osteoarthritis: A Randomised Clinical Trial

**DOI:** 10.1101/2025.09.19.25336135

**Authors:** Md. Nazmul Huda, Md. Obaidul Haque, Polok Halder, Nadia Afrin Urme

**Affiliations:** Department of Physiotherapy, Bangladesh Health Professions Institute, CRP, Savar, Dhaka - 1343, Bangladesh; Department of Physiotherapy and Rehabilitation, Jashore University of Science & Technology, Jashore-7408, Bangladesh; Centre for the Rehabilitation of the Paralysed (CRP) – Savar, Chapain, Savar, Dhaka- 1343, Bangladesh

**Keywords:** Knee osteoarthritis, Mobilisation with Movement, physiotherapy, randomised clinical trial

## Abstract

**Introduction:** Knee osteoarthritis (OA) is a degenerative joint disorder involving progressive cartilage loss, subchondral bone changes, and periarticular tissue involvement. It is a major cause of pain, stiffness, and reduced mobility, particularly in weight-bearing joints. Mobilisation with Movement (MWM), a Mulligan Concept manual therapy, aims to correct minor joint positional faults during active movement, potentially reducing pain and restoring function. This study examined the effectiveness of MWM with usual physiotherapy care compared with usual care alone in individuals with knee OA.

**Methods and analysis:** A single-blind randomised controlled trial was conducted from June to August 2025 at the Musculoskeletal Physiotherapy Unit, Centre for the Rehabilitation of the Paralysed (CRP), Bangladesh. Fifty adults aged 40–65 years with radiographically confirmed knee OA (Kellgren–Lawrence grade ≥2) were allocated to MWM plus usual care (n=25) or usual care alone (n=25). Both groups received 30-minute sessions, three times weekly for six weeks. Outcomes included pain (Numeric Pain Rating Scale, NPRS), active and passive knee range of motion (ROM), and physical function (Western Ontario and McMaster Universities Osteoarthritis Index, WOMAC). The primary outcome was pain intensity (NPRS). Secondary outcomes included knee ROM and WOMAC scores.

**Results:** Both groups improved significantly (p<0.001), but the MWM group achieved greater pain reduction (median NPRS change –4.0 vs –2.0; p<0.001), larger ROM gains, and greater WOMAC score improvements (mean change –19.2 vs –10.8; p<0.001).

**Conclusions:** MWM combined with usual care produced superior improvements in pain, ROM, and function versus usual care alone, supporting its integration into physiotherapy protocols for knee OA.

**Clinical Trial Registry India:** CTRI/2025/05/086343 [Registered on: 05/05/2025]

## INTRODUCTION

Knee osteoarthritis (OA) is one of the most prevalent musculoskeletal disorders, causing pain, stiffness, and functional disability worldwide. It results from progressive degeneration of articular cartilage, alterations in subchondral bone, and inflammation of periarticular structures, ultimately leading to impaired mobility and reduced quality of life [1, 2]. The knee joint, due to its weight-bearing role and limited inherent stability, is particularly vulnerable to OA-related structural and symptomatic decline [3].

The global prevalence of knee OA is estimated to be around 16%, with women and older adults disproportionately affected [4]. South Asian populations report variable but high rates, ranging from 22% to 39% in India and 21.6% in Pakistan, whereas the prevalence in Bangladesh has been reported as 7.7% [5, 6]. Risk factors include age, obesity, genetic predisposition, prior injury, and sedentary lifestyles [7]. With increasing life expectancy and lifestyle changes, the incidence and burden of knee OA are projected to rise further, intensifying its impact on health systems.

Conservative management remains the first-line approach and typically involves exercise therapy, weight management, pharmacological agents, and physiotherapy modalities. Surgical procedures such as total knee arthroplasty are considered in advanced cases but are costly, invasive, and not universally accessible [8]. Within physiotherapy, manual therapy techniques have received increasing attention as non-invasive strategies for symptom relief and functional restoration.

Mobilisation with Movement (MWM), developed by Brian Mulligan, involves therapist-applied sustained glides combined with the patient’s active movement. This approach is proposed to correct subtle joint positional faults and has been associated with reductions in pain, improvements in range of motion (ROM), and enhanced functional outcomes [9, 10]. While preliminary studies report favorable results, high-quality randomised trials comparing MWM with conventional physiotherapy in knee OA remain limited, particularly in resource-constrained settings.

Therefore, this trial aimed to evaluate the effectiveness of MWM combined with usual physiotherapy care compared with usual care alone in individuals with knee OA, focusing on outcomes related to pain, ROM, and physical function.

## METHODS

### Study design

This trial was designed as a single-blind, parallel-group, randomised controlled study conducted between June and August 2025. The objective was to compare the clinical effects of Mobilisation with Movement (MWM) in combination with usual physiotherapy care against usual care alone in patients diagnosed with knee osteoarthritis. Participants were randomly assigned in equal numbers to the experimental or control arm using a computer-generated randomisation procedure. Blinding was maintained for participants and outcome assessors. The trial was prospectively registered in the Clinical Trial Registry of India (CTRI/2025/05/086343) and followed the CONSORT (Consolidated Standards of Reporting Trials) statement to ensure transparency and reproducibility (Figure 1).

**Figure 1:**
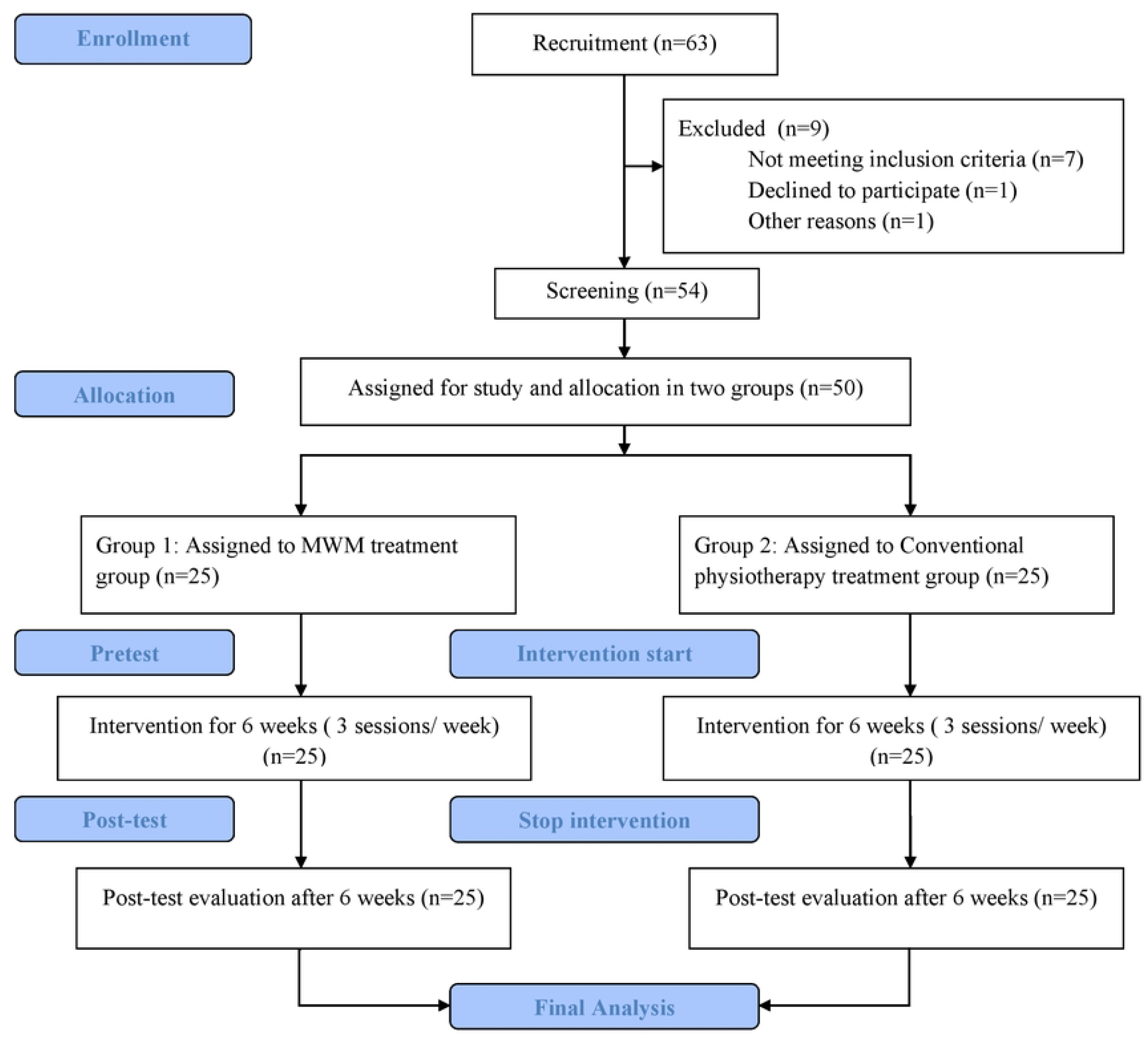
Reporting Guideline [Consolidated Standards for Reporting Trials (CONSORT)]

### Study setting

The study was carried out in the Musculoskeletal Physiotherapy Outpatient Department of the Centre for the Rehabilitation of the Paralysed (CRP), Savar, Dhaka, Bangladesh. CRP is a national tertiary centre for rehabilitation, receiving referrals from across the country. Its musculoskeletal unit provides a high caseload of patients with knee osteoarthritis from diverse socio-economic backgrounds, making it an appropriate and representative setting for clinical trials.

### Study population

The target population included adult men and women with symptomatic knee osteoarthritis who attended the musculoskeletal outpatient services of CRP during the study period. The sample reflected a heterogeneous group representing both urban and rural communities, thereby capturing the demographic diversity of individuals affected by knee osteoarthritis in Bangladesh.

### Sample size calculation

The required sample size was determined using a two-tailed hypothesis test with a 95% confidence level (α = 0.05) and a statistical power of 80% (β = 0.20). Based on an anticipated effect size of 8 and a standard deviation of 10, it was estimated that a minimum of 25 participants per group would be necessary. As a result, a total of 50 individuals were recruited and evenly allocated to the experimental and control groups.

### Recruitment and screening procedures

Between June 1 and June 30, 2025, patients attending the Musculoskeletal Physiotherapy Outpatient Unit at the Centre for the Rehabilitation of the Paralysed (CRP), Savar, Dhaka, were screened for eligibility. A research assistant applied the predefined inclusion and exclusion criteria, and eligible individuals were informed about the study and invited to participate. Written informed consent was obtained before enrollment. Blinded assessors performed baseline demographic and clinical assessments. Following assessment, participants were randomly assigned to either the intervention group (MWM plus usual care) or the control group (usual care only) using a computer-generated concealed allocation process. Randomisation details were placed in sealed, opaque envelopes to maintain allocation concealment. Participants and outcome assessors remained blinded to group assignments. Each participant then completed a 6-week intervention phase, and post-intervention assessments were conducted immediately after treatment. All final assessments were completed by August 25, 2025. No additional follow-up was performed beyond the intervention period.

### Eligibility criteria

A structured screening procedure was applied to determine whether potential participants met the study requirements.

#### Inclusion criteria

Participants were considered eligible if they (1) had a confirmed clinical and radiographic diagnosis of knee osteoarthritis; (2) were aged between 40 and 65 years [11]; (3) included both men and women [12]; (4) had unilateral or bilateral knee osteoarthritis classified as Kellgren– Lawrence grade 2 or higher [13]; and (5) were receiving physiotherapy services at the Musculoskeletal Physiotherapy Unit of CRP.

#### Exclusion criteria

Participants were excluded if they (1) had undergone surgery of the knee or lower limb; (2) had received intra-articular corticosteroid or hyaluronic acid injections within the previous six months [12]; (3) had neurological conditions affecting lower limb function; or (4) reported altered sensation to heat, cold, or pressure around the affected knee [12].

### Sampling technique

A total of 50 patients with knee osteoarthritis who met the eligibility criteria were recruited from the Musculoskeletal Physiotherapy Outpatient Unit at CRP using a simple random sampling approach. After enrolment, participants were allocated equally into two groups: 25 to the experimental arm (MWM plus usual physiotherapy care) and 25 to the control arm (usual care only). Randomisation was carried out with a computer-generated sequence of numbers ranging from 1 to 50. Group assignments were coded as “E” for the experimental group and “C” for the control group, followed by individual participant numbers (e.g., E1, E2… and C1, C2…). This randomisation method was selected to strengthen the internal validity of the study and minimize allocation bias.

### Blinding

The study followed a single-blind design. Participants and outcome assessors were blinded to treatment allocation, while treating physiotherapists could not be blinded due to the nature of the interventions. To preserve blinding, participants were not informed about the intervention provided to the opposite group and were unaware of the specific study hypothesis. Separate assessors were assigned to each group, and treatment sessions were conducted in different rooms on separate floors. Allocation concealment was maintained using sealed opaque envelopes, prepared and handled by an independent researcher not otherwise involved in the trial. Oversight was provided by an independent monitoring team and a trial manager to ensure adherence to blinding procedures throughout the study.

### Details of the interventions

All treatments were delivered by a licensed physiotherapist. Each participant received 30-minute therapy sessions, conducted three times per week over six weeks.

#### Intervention group

Patients assigned to the intervention group underwent Mobilisation with Movement (MWM). This involved sustained manual glides of the tibia in different planes—medial, lateral, anterior, posterior, or rotational—while the participant actively flexed and extended the knee. The procedures were guided by the principles described in Mulligan’s manual therapy text (Mulligan, 2004). Each patient was first assessed in a supine position, where glides were systematically tested across the frontal, sagittal, and rotational planes. The glide direction that provided the greatest pain relief and range-of-motion gain was selected for treatment. When pain-free motion was achieved, gentle end-range overpressure was applied. If the participant remained symptom-free in supine testing, glides were further evaluated in weight-bearing positions, and the most effective direction was then adopted. Treatment consisted of 10 repetitions, performed in three sets per session. Sessions were provided three times weekly for six consecutive weeks.

- Manual Glide – Non-Weight-Bearing – Flexion and Extension
- Lateral glide MWM for flexion (6–10 repetitions × 3–5 sets)
- Lateral glide MWM for extension (6–10 repetitions × 3–5 sets)
- Medial glide MWM for flexion (6–10 repetitions × 3–5 sets)
- Medial glide MWM for extension (6–10 repetitions × 3–5 sets)

#### Control group

Participants in the control arm received only conventional physiotherapy interventions. These included:

- Stretching exercise: 15–30 seconds hold with 3–5 repetitions, 3 sets
- Isometric exercise: 10 seconds of contraction with 10 repetitions, 3 sets
- Eccentric exercise: 10 repetitions, 3 sets
- Maitland mobilisation: 10 repetitions, 3 sets
- Infrared radiation (IRR) or Ice therapy: 10 minutes

### Method of Data Collection

#### Measurement tools

Data collection was performed using validated assessment tools to evaluate pain, joint mobility, and functional status. Three primary instruments were employed: the Numeric Pain Rating Scale (NPRS), a universal goniometer, and the Western Ontario and McMaster Universities Osteoarthritis Index (WOMAC). Sociodemographic information was also collected at baseline using a structured questionnaire.

#### Numeric Pain Rating Scale (NPRS)

Pain intensity was assessed using the NPRS, a widely used self-report measure. Participants were asked to rate their pain on a scale from 0 to 10, where 0 indicated “no pain” and 10 represented “worst imaginable pain.” The NPRS is easy to administer and interpret, making it a practical tool for evaluating subjective pain levels in clinical and research settings. While it lacks detailed information on pain characteristics, its simplicity and responsiveness make it suitable for tracking treatment-related changes over time [14].

#### Goniometric assessment

Knee range of motion (ROM) was measured using a standard universal goniometer. Active and passive flexion and extension were assessed with participants in the supine position. Each direction was tested individually, and the corresponding angle was recorded for analysis. Goniometric measurements have demonstrated acceptable inter-rater reliability in clinical studies, with intraclass correlation coefficients (ICCs) typically ranging from 0.59 to 0.90 [15].

#### Western Ontario and McMaster Universities Osteoarthritis Index (WOMAC)

Functional ability was assessed using the WOMAC questionnaire, which evaluates pain, stiffness, and physical function. The scale uses a five-point Likert system, with responses ranging from 0 (none) to 4 (extreme difficulty). Participants completed the questionnaire independently. WOMAC has been shown to have good reliability and validity for individuals with hip or knee osteoarthritis, and is commonly used in clinical trials assessing treatment outcomes [16].

### Study procedure and data collection methods

The study followed a structured process involving pre-intervention assessment, a six-week treatment phase, and post-intervention evaluation. Each participant received 18 physiotherapy sessions over six weeks, supervised by a trained physiotherapist who also conducted the assessments. At baseline, participants completed a structured questionnaire capturing sociodemographic information. Pain intensity was measured using the Numeric Pain Rating Scale (NPRS), knee range of motion (ROM) was assessed with a universal goniometer, and functional status was evaluated using the Western Ontario and McMaster Universities Osteoarthritis Index (WOMAC). Following the final treatment session, the same tools and procedures were used to reassess outcomes. The physiotherapist recorded the objective measures (e.g., ROM), while participants completed the subjective components (e.g., NPRS and WOMAC). All data were documented on predesigned case report forms. Both the experimental and control groups were assessed under identical conditions to ensure uniformity and reduce bias. This standardised approach allowed for consistent, reliable data collection across the trial period.

### Data management

To ensure data accuracy and integrity, all records were reviewed regularly for errors and omissions. Assessors verified the data daily after each evaluation. Manual data entry was performed twice to minimise transcription errors. The final dataset was accessible to the trial manager, lead investigator, and designated data auditors. Each participant was assigned a unique identification code to anonymise personal information. Only the lead investigator retained access to both hard copies and digital files, which were stored securely within the physiotherapy postgraduate research unit on password-protected servers. A master list linking participant IDs to identities was stored separately for security purposes. De-identified data were used for statistical analysis, and all findings were reported in aggregated form to preserve participant confidentiality. Post-trial care was considered only in cases where adverse effects were observed during the study.

### Statistical analysis

Data analysis was performed using SPSS version 25.0 (IBM Corp., Armonk, NY) and Microsoft Excel 2016. All entries were double-checked for completeness and accuracy before analysis. Analyses followed an intention-to-treat approach. Continuous variables were checked for normality using the Shapiro–Wilk test. The primary outcome was the change in pain intensity (NPRS) from baseline to 6 weeks. Because NPRS data were not normally distributed, within-group changes were assessed using the Wilcoxon signed-rank test, and between-group comparisons were performed using the Mann–Whitney U test. For the secondary outcomes (knee ROM and WOMAC scores), variables were approximately normally distributed; therefore, within-group changes were analyzed using paired t-tests, and between-group differences were examined using independent t-tests. All tests were two-tailed, and a p-value <0.05 was considered statistically significant. No imputation was performed for missing data.

### Monitoring

A monitoring team consisting of two independent members, neither of whom was involved in the treatment or assessment phases, was appointed to ensure the study’s procedural integrity. Their responsibilities included observing the delivery of interventions, tracking participant enrolment, and monitoring for adverse events. They also reviewed the data and conducted preliminary oversight of the analytical process. Any protocol modifications or procedural changes were reported to the Institutional Review Board (IRB) by the principal investigator for approval.

### Safety measures and adverse effects management

Although significant side effects were not anticipated from the physiotherapy interventions, the monitoring team closely observed for any unexpected events during and after the treatment sessions. Any adverse occurrences were promptly reported to the principal investigator and documented in the treating therapist’s SOAP notes. Participants were advised to notify their physiotherapist immediately if they experienced pain, discomfort, or skin irritation. Should any serious adverse effects have occurred, they would have been reported in the final trial manuscript and reviewed by the monitoring team. Any necessary protocol adjustments or safety measures were to be communicated to the IRB without delay.

### Ethical considerations and informed consent

Ethical approval for this study was obtained from the Institutional Review Board of the Bangladesh Health Professions Institute (Approval No. BHPI/IRB/10/2023/796). This trial was prospectively registered with the Clinical Trial Registry of India (CTRI/2025/05/086343) on May 5, 2025, before the enrolment of the first participant. All procedures adhered to the ethical principles of the Declaration of Helsinki. Written informed consent was obtained from all participants before enrolment. Participation was entirely voluntary, and individuals were informed of their right to withdraw at any point without any consequence to their clinical care. Only authorised personnel had access to anonymised data, all of whom were bound by confidentiality agreements. In the event of any adverse outcomes, appropriate post-trial care was to be provided. The authors confirm that all ongoing and related trials for this intervention are registered.

### Patient and public involvement

Patients and members of the public were not involved in the design, conduct, reporting, or dissemination of this research.

## RESULTS

### Baseline Characteristics

The baseline characteristics of both groups are presented in Table 1. Participants in the control group had a mean age of 51.40 years, whereas the mean age in the experimental group was 53.00 years, indicating both groups were middle-aged and comparable. The average pain score was slightly higher in the experimental group (7.20 ± 1.19) compared to the control (6.76 ± 1.64). Active flexion of the knee was identical in both groups (113.60°). Active extension loss was marginally greater in the experimental group (–13.60 ± 3.95) compared with the control (–12.80 ± 3.25). Passive flexion showed nearly similar values across groups, while passive extension loss followed a comparable pattern. WOMAC scores were somewhat higher in the experimental group (54.95 ± 9.04) than in the control (41.66–70.83). These findings suggest the groups were overall well-balanced at baseline, allowing valid comparison (Table 1).

**Table 1:** Baseline Characteristics of Participants in Experimental and Control Groups.

| Variable | Experimental (Mean ± SD) | Control (Mean ± SD) |
| --- | --- | --- |
| Age (years) | $51.40 \pm 10.89$ | $53.00 \pm 9.86$ |
| Pain (Numeric Pain Rating Scale) | $7.20 \pm 1.19$ | $6.76 \pm 1.64$ |
| Active Knee Flexion (degrees) | $113.60 \pm 8.60$ | $113.60 \pm 8.60$ |
| Active Knee Extension (degrees) | -13.60 ± 3.95 | -12.80 ± 3.25 |
| Passive Knee Flexion (degrees) | 116.20 ± 11.20 | 118.20 ± 8.77 |
| Passive Knee Extension (degrees) | -8.40 ± 4.01 | -7.80 ± 3.25 |
| WOMAC Score (Western Ontario and McMaster Universities Arthritis Index) | 54.95 ± 9.04 | 41.66–70.83 |

### Socio-Demographic Information

Socio-demographic characteristics are summarized in Table 2. The highest proportion of participants in both groups was between 51 and 60 years of age. More women than men participated, with women representing 52% of the control group and 64% of the experimental group. Most participants were married, from semi-urban or rural areas, and had completed secondary education. More than half of the participants reported a monthly income below 10,000 BDT. Family type distribution also varied, with extended families reported more frequently in the control group, while nuclear families were more common in the experimental group (Table 2).

**Table 2:** Demographic and Socioeconomic Characteristics of Participants.

| Variable | Control Group (n=25), n (%) | Experimental Group (n=25), n (%) |
| --- | --- | --- |
| Age Category (years) | 21–30: 2 (8%) | 21–30: 1 (4%) |
|  | 31–40: 3 (12%) | 31–40: 3 (12%) |
|  | 41–50: 4 (16%) | 41–50: 9 (36%) |
|  | 51–60: 13 (52%) | 51–60: 7 (28%) |
|  | 61–70: 3 (12%) | 61–70: 5 (20%) |
| Gender | Women: 13 (52%)<br>Men: 12 (48%) | Women: 16 (64%)<br>Men: 9 (36%) |
| Area of Living | Urban: 4 (16%)<br>Semi-urban: 9 (36%)<br>Rural: 12 (48%) | Urban: 5 (20%)<br>Semi-urban: 9 (36%)<br>Rural: 11 (44%) |
| Educational Level | Illiterate: 3 (12%)<br>Secondary: 15 (60%)<br>H.S.C.: 3 (12%)<br>Graduation & above: 4 (16%) | Illiterate: 8 (32%)<br>Secondary: 13 (52%)<br>H.S.C.: 3 (12%)<br>Graduation & above: 1 (4%) |
| Monthly Income (Taka) | 0–10,000: 13 (52%)<br>11,000–20,000: 6 (24%)<br>21,000–30,000: 2 (8%)<br>31,000–40,000: 3 (12%)<br>41,000–50,000: 1 (4%) | 0–10,000: 16 (64%)<br>11,000–20,000: 4 (16%)<br>21,000–30,000: 4 (16%)<br>41,000–50,000: 1 (4%) |
| Marital Status | Married: 24 (96%)<br>Single: 1 (4%) | Married: 25 (100%) |
| Family Type | Nuclear: 12 (48%)<br>Extended: 13 (52%) | Nuclear: 15 (60%)<br>Extended: 10 (40%) |

### Pain (Numeric Pain Rating Scale, NPRS)

Analysis of pain scores indicated significant improvement following treatment in both groups. Within-group analysis revealed that both the control and experimental groups demonstrated a highly significant reduction in pain after intervention (Z = 4.667, p = 0.001 for both) (Table 3). When groups were compared, a significant difference emerged in favor of the experimental group, with the Mann–Whitney U test confirming superior pain reduction with MWM (U = 4.792, p = 0.001) (Table 4).

**Table 3:** Within-Group Comparison of Pain Scores (NPRS) Using Wilcoxon Signed Rank Test.

| Group | Sample Size (n) | Z-value | p-value |
| --- | --- | --- | --- |
| Control | 24 | 4.667 | 0.001* |
| Experimental | 24 | 4.667 | 0.001* |

\*Significant at $p < 0.05$

**Table 4:** Between-Group Comparison of Pain Scores (NPRS) Using Mann-Whitney U Test.

| Variable | df | U-value | p-value |
| --- | --- | --- | --- |
| NPRS | 48 | 4.792 | 0.001* |

\*Significant at $p < 0.05$

### Range of Motion (ROM)

Changes in knee ROM are shown in Tables 5 and 6. Within-group analyses demonstrated statistically significant improvements in both active flexion, active extension, passive flexion, and passive extension in the two groups following treatment (all p ≤ 0.001). Between-group comparisons revealed that the experimental group achieved significantly greater improvements in active flexion (p = 0.001) and passive flexion (p = 0.001) compared with the control. However, there were no statistically significant differences between groups for active extension (p = 0.188) and passive extension (p = 0.776) (Table 6).

**Table 5:** Within-Group Changes in Knee Range of Motion (Paired t-test)

| Group | ROM Measure | df | t-value | p-value |
| --- | --- | --- | --- | --- |
| Control | Active Flexion | 24 | 10.830 | 0.001* |
| Control | Active Extension | 24 | 15.501 | 0.001* |
| Control | Passive Flexion | 24 | 10.864 | 0.001* |
| Control | Passive Extension | 24 | 15.501 | 0.001* |
| Experimental | Active Flexion | 24 | 8.697 | 0.001* |
| Experimental | Active Extension | 24 | 11.988 | 0.001* |
| Experimental | Passive Flexion | 24 | 7.918 | 0.001* |
| Experimental | Passive Extension | 24 | 11.854 | 0.001* |

\*Significant at $p < 0.05$

**Table 6:**
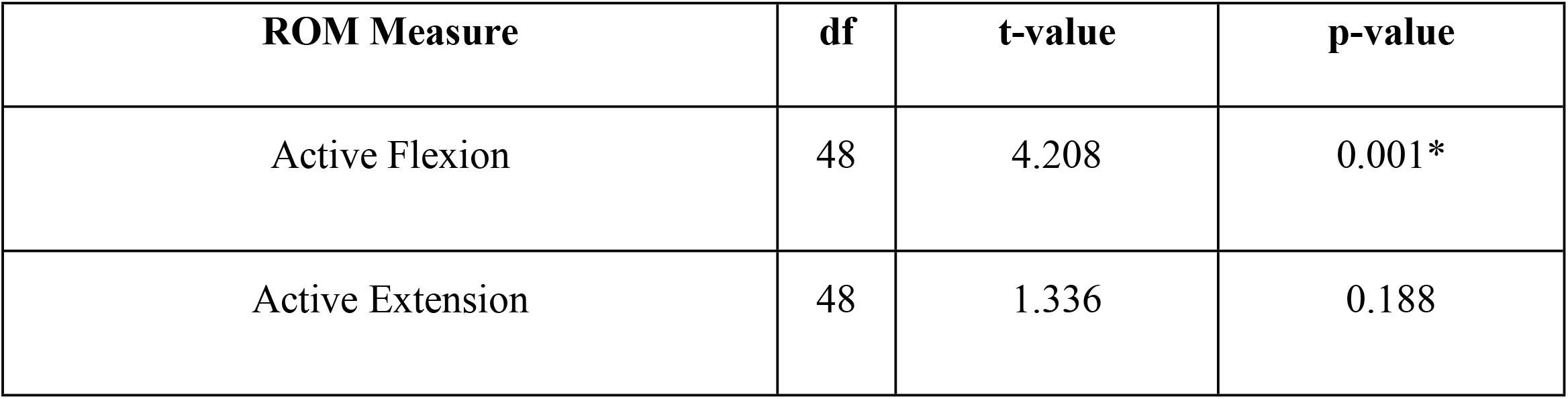
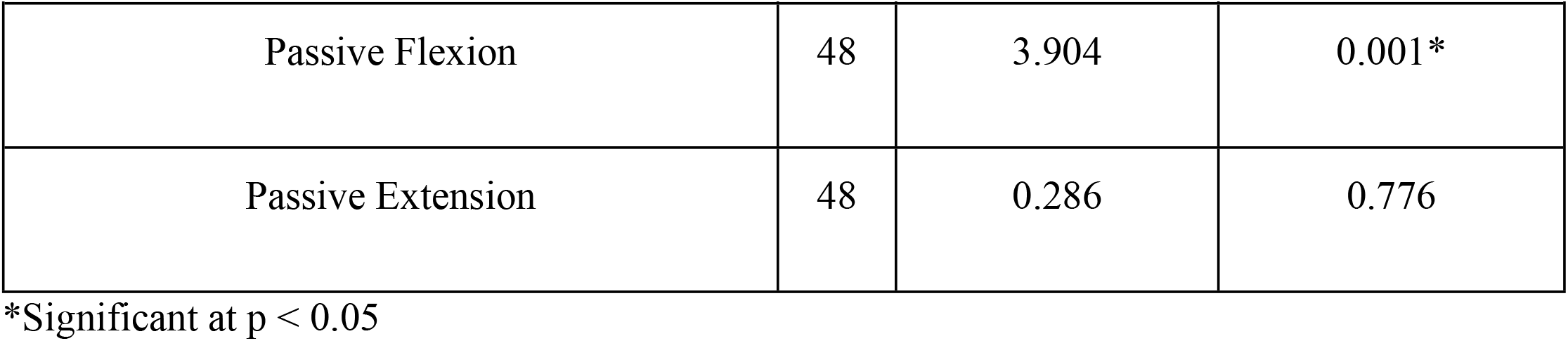
Between-Group Comparison of Knee Range of Motion (Independent t-test)

### Functionality (WOMAC Index)

Functional outcomes measured using the WOMAC scale are presented in Tables 7 and 8. Within-group comparisons showed highly significant improvements in both the control and experimental groups after intervention (p = 0.001 for each). However, between-group analysis demonstrated greater improvement in the experimental group, with a significant difference favoring MWM (t = 5.562, p = 0.001) (Table 8).

**Table 7:** Within-Group Changes in Functional Ability (WOMAC) Using Paired t-test.

| Group | df | t-value | p-value |
| --- | --- | --- | --- |
| Control | 24 | 13.263 | 0.001* |
| Experimental | 24 | 29.775 | 0.001* |

\*Significant at $p < 0.05$

**Table 8:** Between-Group Comparison of Functional Ability (WOMAC) Using Independent t-test.

| df | t-value | p-value |
| --- | --- | --- |
| 48 | 5.562 | 0.001* |

\*Significant at $p < 0.05$

In summary, both groups showed significant improvements following treatment; however, patients who received Mobilization with Movement achieved superior outcomes in terms of pain reduction, knee flexion range of motion, and functional ability. Differences in knee extension were not statistically significant. These results suggest that the addition of MWM provides greater clinical benefit than usual physiotherapy care in the management of knee osteoarthritis.

## DISCUSSION

This randomized clinical trial demonstrated significant improvements in pain, knee flexion range of motion, and functional ability in patients with knee osteoarthritis (OA) who received Mobilisation with Movement (MWM) combined with usual care compared to usual physiotherapy care alone. These findings align with previous studies that have highlighted the benefits of MWM for musculoskeletal conditions, particularly knee OA.[12, 17, 18] The enhanced pain relief observed may be attributed to the neurophysiological and biomechanical effects of MWM, including improved joint kinematics, reduction of pain facilitatory mechanisms, and increased confidence in movement [19].

Knee flexion range of motion improved significantly more in the MWM group, which is crucial as limited flexion directly impacts performing daily activities like sitting and climbing stairs.[20] However, the between-group difference in knee extension was not statistically significant. This could be due to chronic capsular contractures or structural changes such as osteophytes found in chronic OA, which may restrict extension and respond less readily to manual therapy interventions within the short time frame of this study [21, 22].

Functional improvement measured by WOMAC scores was also greater in the MWM group, reflecting the compounded effect of pain reduction and improved mobility on patients’ quality of life. These results resonate with the findings of the study, which reported that manual therapy combined with exercise enhances overall function more effectively than exercise alone [7].

Despite these promising results, certain limitations need to be acknowledged. First, the sample size was relatively small and drawn from a single geographic region, limiting broader generalizability. Larger multicenter trials would be valuable to confirm these findings across diverse populations. Second, the duration of the intervention and follow-up was limited; longitudinal studies are needed to assess long-term benefits and the sustainability of MWM effects in knee OA.

Additionally, blinding participants and therapists in manual therapy trials is inherently challenging, which could introduce performance bias. To minimize this, objective outcome measures such as range of motion were included alongside subjective pain and function scores. Potential confounding factors like adherence to home exercises, use of concomitant medications, and variations in usual care protocols could have influenced results, although randomization and baseline matching helped to mitigate this.

Lastly, structural joint changes (e.g., radiographic severity) were not incorporated as covariates, which may explain variability in response to treatment and limit the ability to tailor interventions optimally [23].

Given MWM’s non-invasive, cost-effective nature and ease of integration into outpatient physiotherapy settings, especially in resource-limited environments like ours, this study adds valuable evidence supporting its adoption for knee OA management. Education of physiotherapists through workshops and clinical seminars, coupled with the dissemination of these findings in open-access forums, can enhance uptake and improve patient outcomes.

Overall, MWM in conjunction with usual care produces clinically meaningful improvements in pain, knee flexion, and function compared to usual care alone in knee OA patients. Future research with larger samples, longer follow-up, and consideration of joint structural status will strengthen the understanding and application of this manual therapy approach.

## Data Availability

The datasets used and analyzed during the study are available from the corresponding author upon reasonable request.

## Declarations

### Grant information

The trial is self-funded by the authors.

### Competing interests

The authors declared that they have no known competing financial interests or personal relationships that could have appeared to influence the work that will be reported in this paper.

### Author’s contribution

**Md. Nazmul Huda:** Conceptualization, Methodology, Writing - Original Draft, Writing - Review & Editing. **Md. Obaidul Haque:** Writing - Review & Editing, Resources, Validation. **Polok Halder:** Writing - Review & Editing, Investigation, Validation. **Nadia Afrin Urme:** Supervise, perform formal analysis, and conduct data curation.

## Supporting Information captions

Supplementary file 1: IRB Approval

Supplementary file 2: Clinical Trial Registry-India

Supplementary file 3: CONSORT Checklist

Supplementary file 4: Informed Consent

## Notes

### Competing Interest Statement

The authors have declared no competing interest.

### Clinical Trial

The trial was prospectively registered with the Clinical Trial Registry of India (CTRI/2025/05/086343) on May 5, 2025.

### Clinical Protocols

https://bmjopensem.bmj.com/content/11/2/e002735

### Funding Statement

The author(s) received no specific funding for this work.

### Author Declarations

Ethical approval for this study was obtained from the Institutional Review Board of the Bangladesh Health Professions Institute (Approval No. BHPI/IRB/10/2023/796), and the trial was prospectively registered with the Clinical Trial Registry of India (CTRI/2025/05/086343) on May 5, 2025. All procedures adhered to the ethical principles of the Declaration of Helsinki. Written informed consent was obtained from all participants before enrolment. Participation was entirely voluntary, and individuals were informed of their right to withdraw at any point without any consequence to their clinical care. Only authorised personnel had access to anonymised data, all of whom were bound by confidentiality agreements. In the event of any adverse outcomes, appropriate post-trial care was to be provided.

